# Mapping the Landscape of Pediatric Clinical Ethics Consults: A Systematic Scoping Review Protocol

**DOI:** 10.1101/2025.05.15.25327592

**Authors:** Matthew Lin, Emma Tunstall, Charles Schufreider, Gwynne Latimer, Daniel Eison, Catherine Constable, Arthur Caplan, New York University Hassenfeld Children’s Hospital’s Pediatric Bioethics Advisory Group

## Abstract

**Background:** Clinical ethics consultation (CEC) services aim to improve the quality and delivery of healthcare by identifying, evaluating, and resolving ethical questions. Approximately one third of children’s hospitals lack CEC services, although this number has increased over the past decade. While there is robust literature on the characteristics, processes, and outcomes of adult CECs, the extent and nature of literature describing pediatric CECs remains unknown. The objective of this systematic scoping review is to map literature that quantifies and characterizes pediatric CECs.

**Methods:** This review will adhere to the Preferred Reporting Items for Systematic Reviews and Meta-Analyses Extension for Scoping Reviews Guidelines (PRISMA-ScR). Inclusion criteria include studies that quantify and characterize pediatric CECs. We will exclude non-English language publications, nonhuman studies, studies that center on maternal-fetal cases, reviews, dissertations, book chapters, and guideline or consensus statements. PubMed, EMBASE, Web of Science, and CINAHL will be searched using a strategy developed by an experienced research librarian. References will be managed in EndNote X9. Title/abstract screening and full text review will be performed using Covidence. Data will be presented in tabular and narrative form.

**Discussion:** Improved understanding of the characteristics, patterns, and trends in pediatric CECs may identify unique and unmet pediatric ethical needs. Identifying these needs may present opportunities to inform initiatives in pediatric ethics program building, education, and research. As research in pediatric clinical ethics evolves, it is also critical to identify the relative strengths and limitations of existing process and outcome typologies, given that there is no universal standard.

## INTRODUCTION

The goal of a clinical ethics consultation (CEC) service is to improve the quality and delivery of healthcare by identifying, evaluating, and resolving ethical issues, questions, or conflicts.^1^ CEC services may represent one mechanism to fulfill the Joint Commission on Accreditation of Healthcare Organizations (JACHO) requirement for having “a process that allows staff, [patients], and families to address ethical issues or issues prone to conflict.”^2^ Robust literature exists around the characteristics, processes, and outcomes of adult CECs,^3–6^ yet less is known about pediatric CECS.

Although the number of children’s hospitals with CEC services has increased over the last decade, more than 30% of children’s hospitals still lack CEC services,^7,8^ which does not align with national standards and practice guidelines.^2,9,10^ Additionally, Children’s hospitals with CEC services display wide variation in staffing, consultant experience, allocation of professional time and resources, and process measures.^7,8^ Pediatric hospitals with CEC Services have shown only moderate adherence to the American Society of Bioethics and Humanities (ASBH) ethics consultation standards.^11^

In addition to the medicolegal and psychosocial dimensions of pediatric care that are distinct from adults, major normative differences also exist in the ethical approach and content in pediatric CECs.^12^ For instance, lack of child autonomy or emerging autonomy necessitating involvement of parents as surrogate decision-makers in most pediatric CECs is not present to the same extent in adults. Subsequently, adult surrogate decision-making frameworks are underpinned more by respect for autonomy and persons, while pediatric decision-making frameworks are often guided by the best interests standard or the harm principle.^13,14^

Empiric descriptions of pediatric CECs have been limited to descriptions of cases mainly at non-freestanding children’s hospitals,^15–22^ with fewer reports from freestanding children’s hospitals^23–31^ and pediatric cancer centers.^32,33^ There have been no published reviews or review protocols that have aimed to capture and characterize the scope of this literature.

### Study Objectives

The overarching goal of this systematic scoping review is to map the literature that describes key characteristics of pediatric CECs. Specific aims include:

1. Identify, summarize, and characterize literature that quantifies and describes pediatric CECs.
2. Describe the demographic, medical, and hospitalization characteristics of pediatric CECs.
3. Identify and summarize process and outcome measures of pediatric CECs.

### Study Rationale

The extent and nature of literature evaluating pediatric CECs remains unknown. A search of PROSPERO, Open Science Framework, Figshare, BioMed Central Protocols, and BMJ Open failed to yield studies, reviews, or protocols with aims similar to our proposed aims. Through informal searches of PubMed and bibliography review, we preliminarily identified 32 studies that quantified and described pediatric CECs (Table 1). We anticipate additional studies that will likely be captured through a more unified search strategy of PubMed and other databases. Almost all studies were published in the United States (US), with two from Switzerland, and one from the Netherlands. By decade, publications on CECs have been the greatest over the last 4 years: 1987-1989=1 study, 1990-1999=3 studies, 2000-2009=4 studies, 2010-2019=11 studies, 2020-2024=12 studies. Pediatric CEC rates vary from 0.5-45 consults per year.

**Table 1.** Relevant studies of pediatric CECs identified through informal PubMed searches and bibliography review.

| Author (publication date) | Study Period | Setting | Single Center | Type of Study | Center | Consults (consult rate) |
| --- | --- | --- | --- | --- | --- | --- |
| La Puma (1987)<sup>38</sup> | 1985-1986 | Mixed | Yes | Retrospective, Survey | University of Chicago | n=2 (2/year) |
| La Puma (1992)<sup>39</sup> | 1988-1989 | Mixed | No | Retrospective | Lutheran General Hospital, University of Chicago | n=2 (2/year) |
| Orr and Perkin (1994)<sup>22</sup> | 1990-1992 | Mixed | Yes | Retrospective | Loma Linda University Medical Center | n=64 (32/year) |
| Yen and Schneiderman (1999)<sup>15</sup> | 1990-1995 | Pediatric | Yes | Retrospective, Mixed-Methods | San Diego Children's Hospital | n=23 (4.6/year) |
| Mitchell and Truog (2000)<sup>40</sup> | 1985-2000 | Pediatric | Yes | Retrospective | Boston Children's Hospital | n=67 (4.4/year) |
| McGee et al (2001)<sup>16</sup> | 1998-1999 | Mixed | No | Survey | 356 hospitals surveyed from the American Hospital's Association Register (1995) | Mean 8.1±15.3 |
| Fox et al (2007)<sup>41</sup> | 1999-2000 | Mixed | No | Survey | 519 hospitals surveyed from the American Hospital Association Survey Register (1998) | Median 3 (0-300) |
| Opel et al (2009)<sup>23</sup> | 1996-2006 | Pediatric | Yes | Retrospective, Mixed-Methods | Seattle Children's Hospital | n=71 (7.1/year) |
| Streuli et al (2014)<sup>24, a</sup> | 2006-2010 | Pediatric | Yes | Retrospective | University Children Hospital Zurich | n=95 (23.8/year) |
| Johnson et al (2015)<sup>32</sup> | 2000-2011 | Pediatric Cancer | Yes | Retrospective | St. Jude's Cancer Center | n=53 (4.8/year) |
| Henricksen et al (2015)<sup>17</sup> | 1999-2014 | Mixed | Yes | Retrospective, Mixed-Methods | Mayo Clinic | n=64 (4.2/year) |
| Thomas et al (2015)<sup>25</sup> | 2005-2013 | Mixed | Yes | Retrospective | Cleveland Clinic | n=102 (12.8/year) |
| Hardart and Lipson (2016)<sup>18</sup> | 2012 | Mixed | No | Survey | 56 freestanding and non-freestanding pediatric centers from the Children's Hospital Association | Median=8 (1-57) |
| Wasson et al (2016)<sup>42</sup> | 2008-2013 | Mixed | Yes | Retrospective, Mixed Methods | Loyola University Medical Center | n=15 (3/year) |
| Boss et al (2018)<sup>43</sup> | 2017 | Mixed | No | Cross-Sectional | 6 Centers associated with the Chronic Critical Illness Collaborative | n=3 (3y/ear) |
| Carter et al (2018)<sup>26</sup> | 2014 | Pediatric | Yes | Retrospective | Childre's Mercy Kansas City Hospital | n=14 (14/year) |
| Watt et al (2018)<sup>27</sup> | 2010-2013 | Pediatric | Yes | Retrospective | Childre's Hospital Colorado | n=47 (15.6/year) |
| Winter et al (2019)<sup>33</sup> | 2007-2017 | Mixed | Yes | Retrospective | Memorial Sloan-Kettering Cancer Center | n=35 (3.5/year) |
| Muggli et al (2019)<sup>44, a</sup> | 2002-2016 | Mixed | No | Retrospective, mixed-methods | Women's University Hospital Basel, Children's University Hospital Basel | n=7 (0.5/year) |
| Friedman et al (2020)<sup>45</sup> | 2020 | Mixed | Yes | Retrospective | Memorial Sloan-Kettering Cancer Center | n=1 (1/year) |
| Vrouenraets et al (2020)<sup>46, b</sup> | 2013-2015 | Mixed | No | Mixed Methods | Amsterdam University Medical Center, Leiden University | n=37 (18.5/year) |
| Leland et al (2021)<sup>19</sup> | 2008-2017 | Mixed | Yes | Retrospective | Indiana University Health | n=165 (18.3/year) |
| Nathanson et al (2021)<sup>28</sup> | 2013-2018 | Pediatric | Yes | Retrospective | Children's Hospital of Philadelphia | n=245 (45/year) |
| Fox et al (2022)<sup>47</sup> | 2016-2017 | Mixed | No | Retrospective | 79 hospitals surveyed from the American Hospital's Association Register (2015) | Median 3 (0-750) |
| Olszewski et al (2023)<sup>30, c</sup> | 2008-2019 | Pediatric | Yes | Case-Control | Northwestern Lurie Children's Hospital | n=209 (19/year) |
| Bosompim et al (2024)<sup>29</sup> | 2001-2019 | Pediatric | Yes | Retrospective, Mixed-Methods | Akron Children's Hospital | n=74 (4.1/year) |
| Freigeh et al (2024)<sup>21</sup> | 2016-2022 | Pediatric | Yes | Retrospective, Mixed-Methods | University of Michigan | n=48 (8/year) |
| Olszewski et al (2024)<sup>48, c</sup> | 2008-2019 | Pediatric | Yes | Retrospective, Mixed Methods | Northwestern Lurie Children's Hospital | n=209 (19/year) |
| James et al (2024)<sup>31</sup> | 2000-2020 | Pediatric | Yes | Retrospective | Dupont Children's Hospital | n=107 (5.3/year) |
| Lyons et al (2024)<sup>20</sup> | 2016-2020 | Mixed | Yes | Retrospective | University of Michigan | n=46 (11.5/year) |
| Siegel et al (2024)<sup>49</sup> | 2012-2021 | Pediatric | Yes | Retrospective | Boston Children's Hospital | n=225 (25/year) |

<sup>a, b</sup> All studies were conducted in the United States, North America, with the exception of: two studies performed in a) Switzerland, Europe, and one study performed in b) Netherlands, Europe.
<sup>c</sup>Two separate studies that used the same cohort of patients with CECs for different analyses.
Setting refers to the following: Mixed = non-freestanding children's hospitals, Pediatric = freestanding children's hospitals.
Consult rates were calculated by dividing the total number of consults by the study period in years

Improved understanding of the characteristics, patterns, and trends in pediatric CECs may identify unique and unmet pediatric ethical needs. Assessing these needs may present opportunities to inform initiatives in pediatric ethics program building and education, that align with the priorities of the AAP, ASBH, JACHO and AMA.^2,9^ As research in pediatric clinical ethics evolves, it is critical to identify the relative strengths and limitations of existing process and outcome typologies, given that there is no universal standard.

## METHODS

This scoping review will adhere to the Preferred Reporting Items for Systematic Reviews and Meta-Analyses Extension for Scoping Reviews guidelines (PRISMA-ScR) to ensure methodological best practices.^34^ A scoping review methodology was selected given our general aims of identifying and characterizing the types of evidence in a given field and clarifying key concepts and definitions.^35^

### Eligibility Criteria

Our review will include studies that quantify and characterize the number of pediatric CECs that occur in or over a given time period. We define pediatric patients as infants, children, adolescents and young adults under the primary care of a pediatric service. We will include studies that address specific pediatric populations or settings (i.e., outpatient clinic, neonatal intensive care units, children with chronic critical illness). We will exclude non-English language publications, studies where full text cannot be obtained, studies of CECs that center on primarily maternal-fetal cases, nonhuman studies, reviews, dissertations, book chapters, and guidelines and consensus statements.

### Information Sources and Search Protocol

The following databases will be searched: PubMed, Embase, Web of Science, and CINAHL. Our search strategy was developed in consultation with an experienced research librarian at New York University Langone Hospital (Table 2). Search terms were based on two concepts: “Pediatric patients/care” and “ethics consultation.” We catalogued MeSH terms and title/text words of relevant studies (n=32) identified by informal preliminary PubMed searches and subsequent bibliography review (Appendix 1). Common MeSH terms (occurring ≥10 times) related to our concepts described above included: Adolescent (15), Child (20), Child, Preschool (12), Decision Making (12), Ethics Consultation (28), Infant (10), Pediatrics (12). Of note, one study was not indexed in PubMed^18^ and two studies did not have MeSH terms listed.^21,29^

**Table 2.** Electronic Database Searches and search strategy (932 combined and 670 after duplicates removed, 650 after non-English studies removed); OR 911 after non-English studies removed, 650 after duplicates from all databases removed.

| Date of search | Database | Search strategy | Studies retrieved | After removal of non-English Studies |
| --- | --- | --- | --- | --- |
| 12/19/2024 | PubMed | ("adolescent"[MeSH Terms] OR "child"[MeSH Terms] OR "child, preschool"[MeSH Terms] OR "infant"[MeSH Terms] OR "pediatrics"[MeSH Terms] OR "pediatric"[All Fields] OR "adolescent"[All Fields] OR "child"[All Fields]) AND ("ethics consult*"[All Fields]) | 271 | 267 |
| 12/19/2024 | CINAHL | (SU (pediatrics or pediatric or adolescent or child) OR AB (pediatrics or pediatric or adolescent or child)) AND TX ethics consult* | 228 | 227 |
| 12/19/2024 | Web of Science | AB=(ethics consult*) AND AB=((pediatrics or pediatric or adolescent or child)) | 242 | 231 |
| 12/19/2024 | EMBASE | (exp pediatrics/ or exp adolescent/ or exp child/ or ("pediatrics" or "pediatric" or "adolescent" or "child").mp.) and ethics consult\$.mp. | 191 | 186 |

Our search strategy as of December 19, 2024, yielded a total of 932 articles from the four databases (Table 2). Duplicate removal yielded 670 title/abstracts and further removal of non-English studies left us with 650 title/abstracts for screening.

### Study Selection

EndNote X9 software will be used for citation collation and duplicates will be removed manually.^36^ Covidence hosted by NYU Langone Health will be used for title and abstract screening followed by full text review by two independent reviewers (ML and GL).^37^ Unresolvable disagreements will be resolved by a third reviewer (CC). Search results will be mapped using the PRISMA-ScR flow diagram.^34^

### Data Charting Process and Data Items

Publications that meet inclusion criteria will undergo full text review and data extraction by two independent reviewers (ML and GL). Extracted content will include the following preliminary categories and responses (Table 3).

**Table 3.** Preliminary data extraction categories and response categories.

| Data Category | Response Categories |
| --- | --- |
| Author (publication date) | Qualitative |
| Journal name | Qualitative |
| Study period(s) | Year or year-year |
| Institution | Freestanding Pediatric Hospital<br>Freestanding Pediatric Cancer Center<br>Children's Hospital within Adult Hospital<br>Other |
| Setting | Inpatient<br>Outpatient<br>Inpatient/Outpatient<br>Other |
| Inclusion of adult CECs in cohort | Yes/No |
| Single center | Yes/No |
| Center Name(s) | Qualitative |
| Country(ies), Continent(s) | Qualitative |
| Type of study | Retrospective cohort<br>Prospective cohort<br>Randomized controlled trial<br>Case-control<br>Cross-sectional, survey/questionnaire<br>Case series<br>Mixed methods<br>Other |
| Additional notes about study type | Qualitative |
| Study aim(s) | Qualitative |
| Inclusion criteria | Qualitative |
| Exclusion criteria | Qualitative |
| Main outcomes addressing study aim(s) | Qualitative, summarize and describe outcomes studied |
| Author Profession | Physician, Non-Physician: Describe |
| Funding received for Study | Yes/No: Public, Private, Other |
| Pediatric CEC number and rate | Total consults (n), total discrete consults if reported (n, i.e., excludes repeat or follow-up consultations)<br>Mean/SD, median/range if reported<br>Consult rate = total consults/study period |
| Statistical analysis of CEC numbers and rates over time | Yes/No, findings |
| Demographics of patients | Race<br>Ethnicity<br>Age<br>Sex<br>Religion<br>Insurance status |
| Medical characteristics of patients | Primary diagnosis/medical problem |
| Structure of CEC Service | Staffing experience/training<br>Resources and support (protected time and financial)<br>Process for accessing/consulting<br>Documentation practices<br>Internal quality and improvement processes |
| CEC core problems or content domains | Describes primary reason, question, or issue that prompted CEC: Yes/No<br>Qualitative analysis of taxonomies |
| CEC analytic frameworks used in analysis | Describes analytic framework used by consultant to addresses question or issue that was posed to CEC service: Yes/No<br>Qualitative analysis of taxonomies |
| CEC process measures | Consult requestor profession: Physician, social work, nursing, patient/family, other<br>Consult requestor department/division<br>Consultation participants in response: 1-on-1 discussion, meeting with clinical team, separate patient/family and team meetings, patient/family/team meetings, other |
| CEC outcome measures as defined by Bell et al<sup>3</sup> | Quality outcomes: Yes/No, qualitative<br>Process outcomes: Yes/No, qualitative<br>Clinical outcomes: Yes/No, qualitative<br>Resource outcomes: Yes/No, qualitative |
| Statistical evaluation of association between demographic, medical, hospitalization, and/or CEC characteristics | Yes/No, qualitative description of findings |

### Critical Appraisal of individual sources of evidence and Risk of Bias

Critical appraisal and bias assessment will not be performed as our aim is to report the breadth of the current literature.

### Synthesis of results

Results will be presented in tabular form to summarize characteristics of included literature, CEC rates, patient demographic, medical, hospitalization and CEC characteristics. Key themes and further knowledge gaps will be discussed in the narrative report.

#### Protocol amendments

Amendments to the protocol will be reported with the results of the review.

## Supporting information

PRISMA-ScR Checklist

## Data Availability

All data produced in the present work are contained in the manuscript

## Acknowledgements

The authors would like to thank Richard McGowan, MLS Head, Education and Clinical Support - NYU Health Sciences Library for instruction in developing our search protocol.

## Author’s contributions

ML conceptualized the study and prepared the draft protocol. GL, CC, DE, and AC critically reviewed and revised the protocol manuscript. All authors read and approved the final manuscript.

## Availability of data and materials

All data generated or analyzed during this study will be included in the published scoping review article

## Ethics approval and consent to participate

This study will not include humans or animals as participants. Data will be sourced from published literature and will therefore not require ethical approval.

## Consent for publication

Not applicable

## Ethics

This study will not include humans or animals as participants. Data will be sourced from published literature and will therefore not require ethical approval from the NYU Grossman School of Medicine Institutional Review Board.

## Abbreviations

AMA: American Medical Association
ASBH: American Society for Bioethics and Humanities
CEC: Clinical Ethics Consultation
PRISMA-ScR: Preferred Reporting Items for Systematic Reviews and Meta-Analyses Extension for Scoping Reviews Guidelines

---

**Appendix 1.**
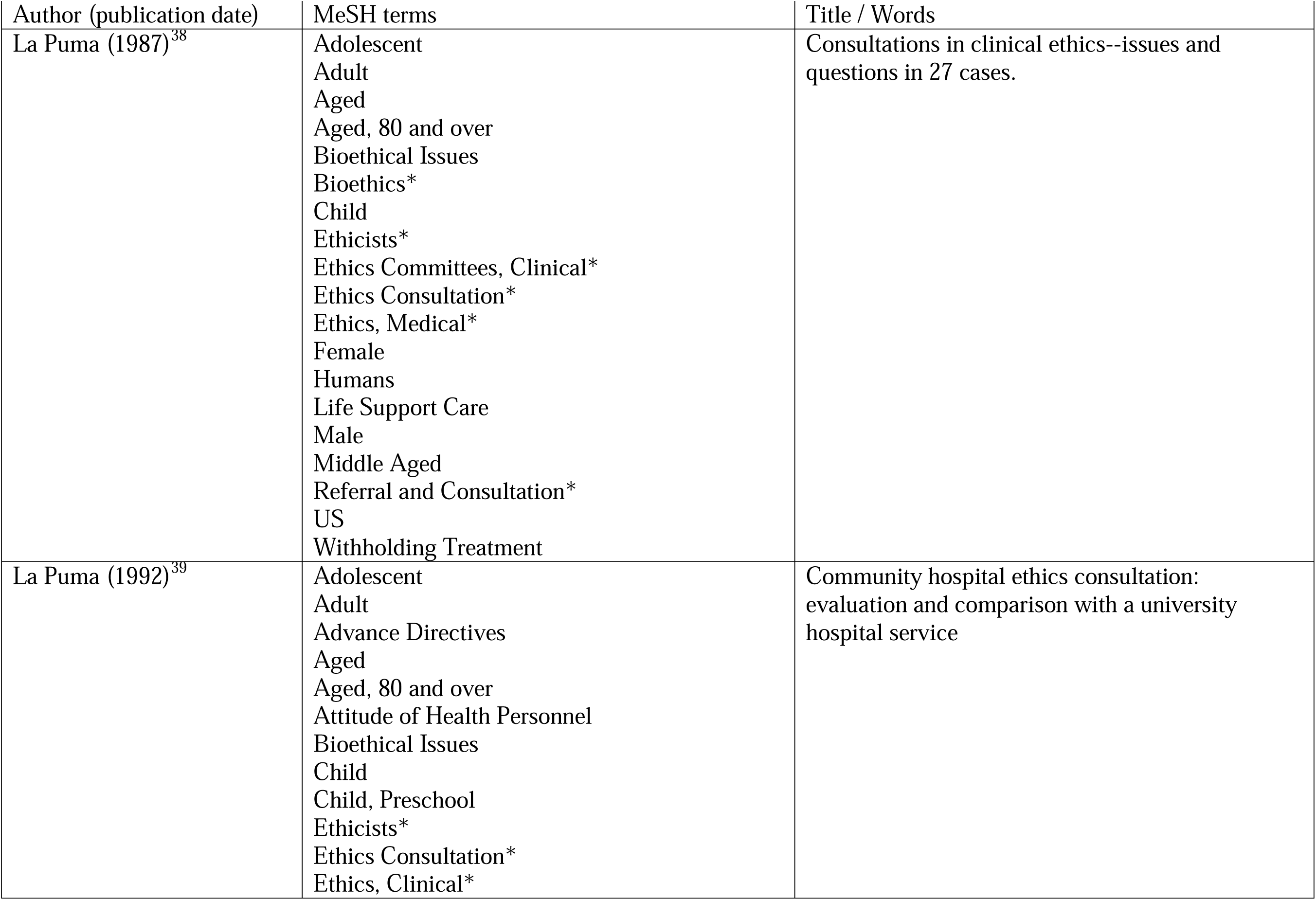
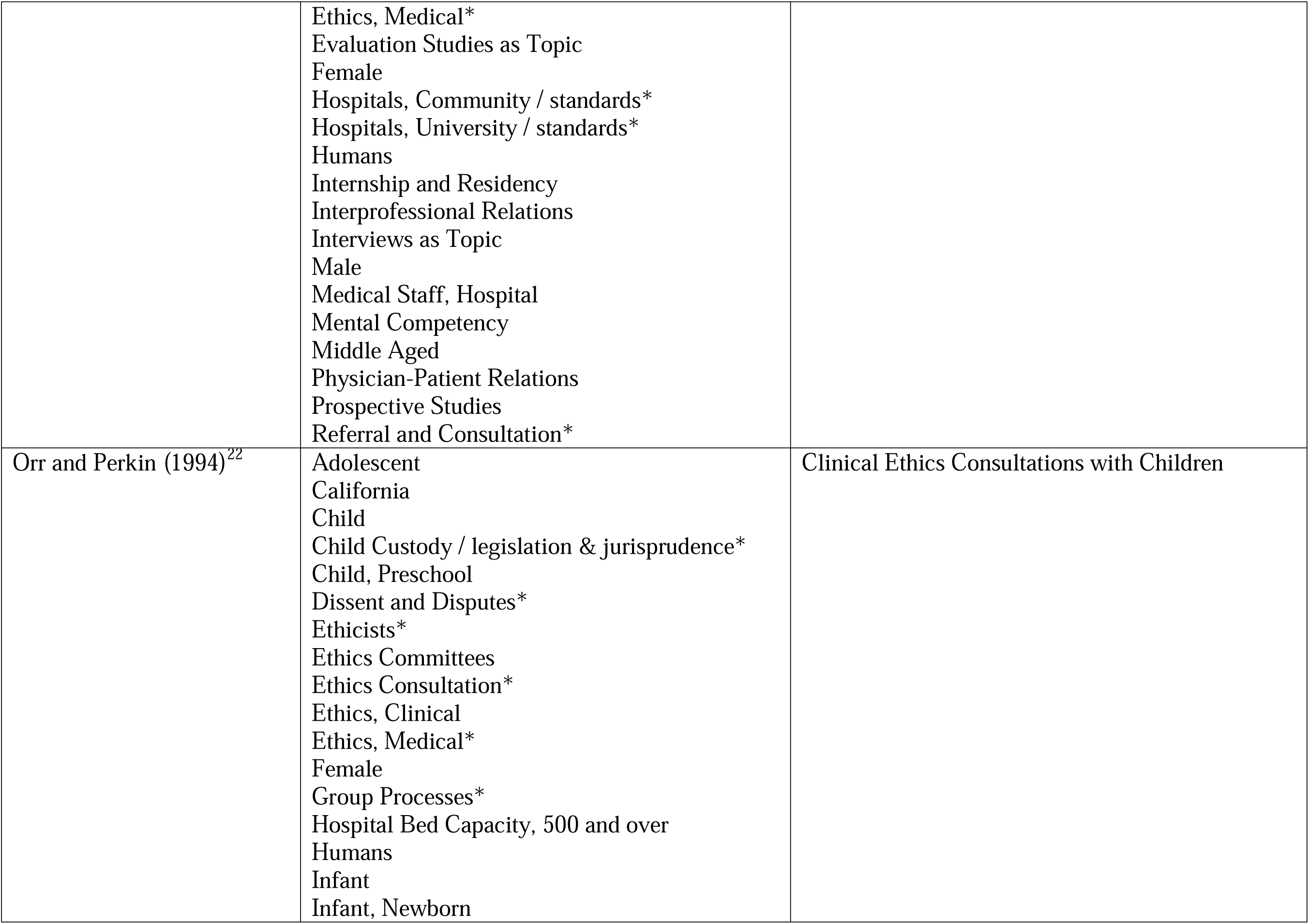
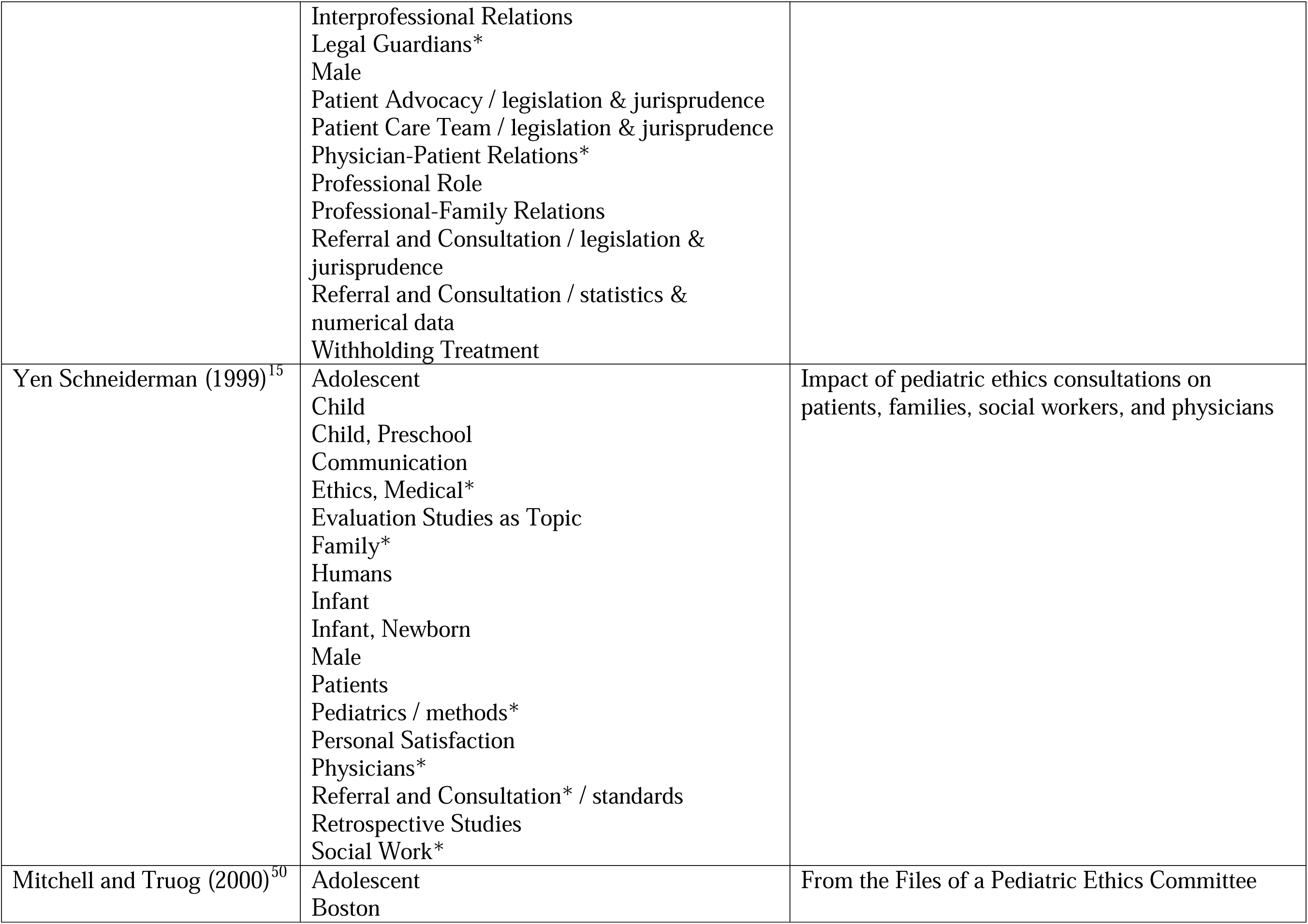
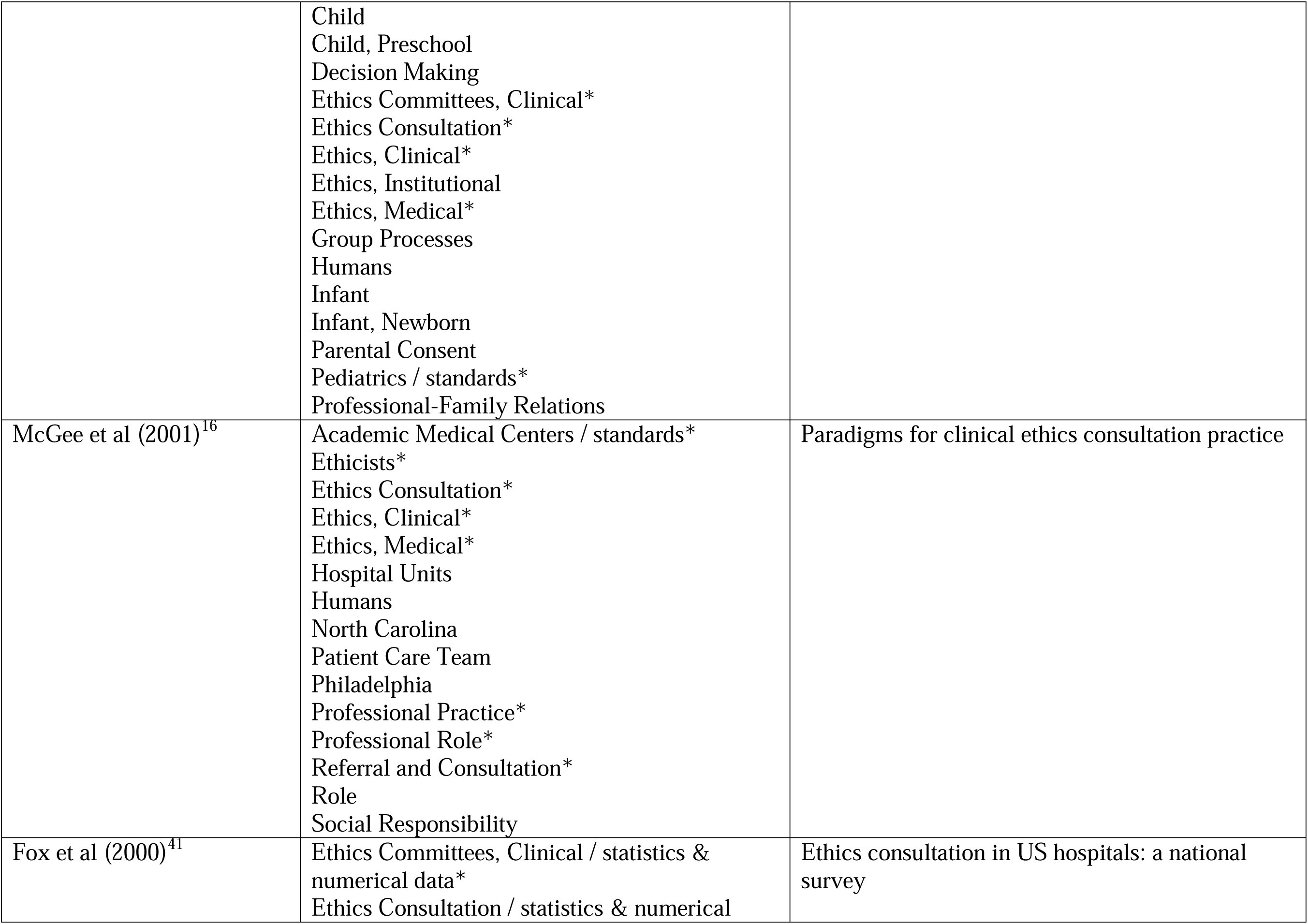
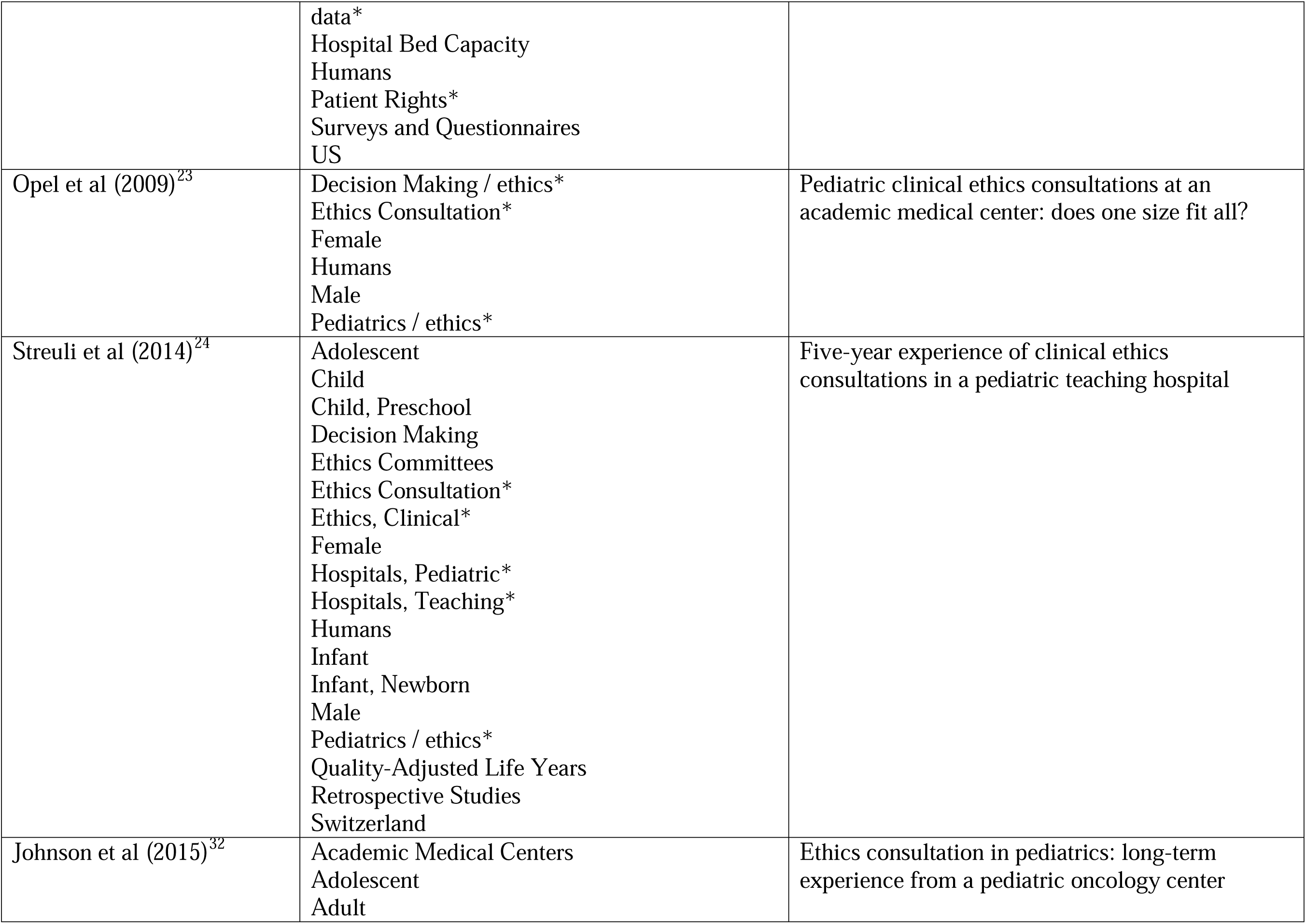
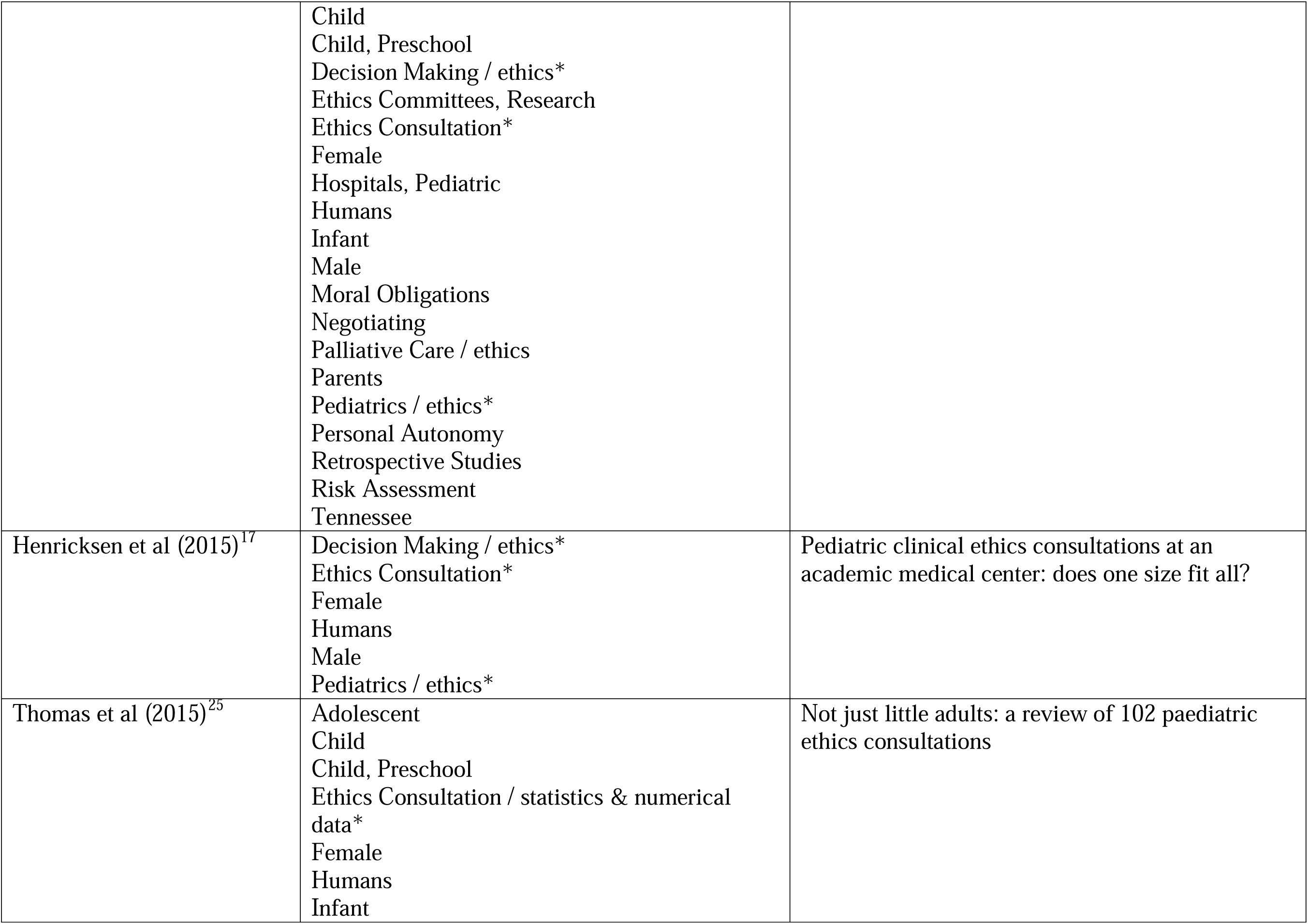
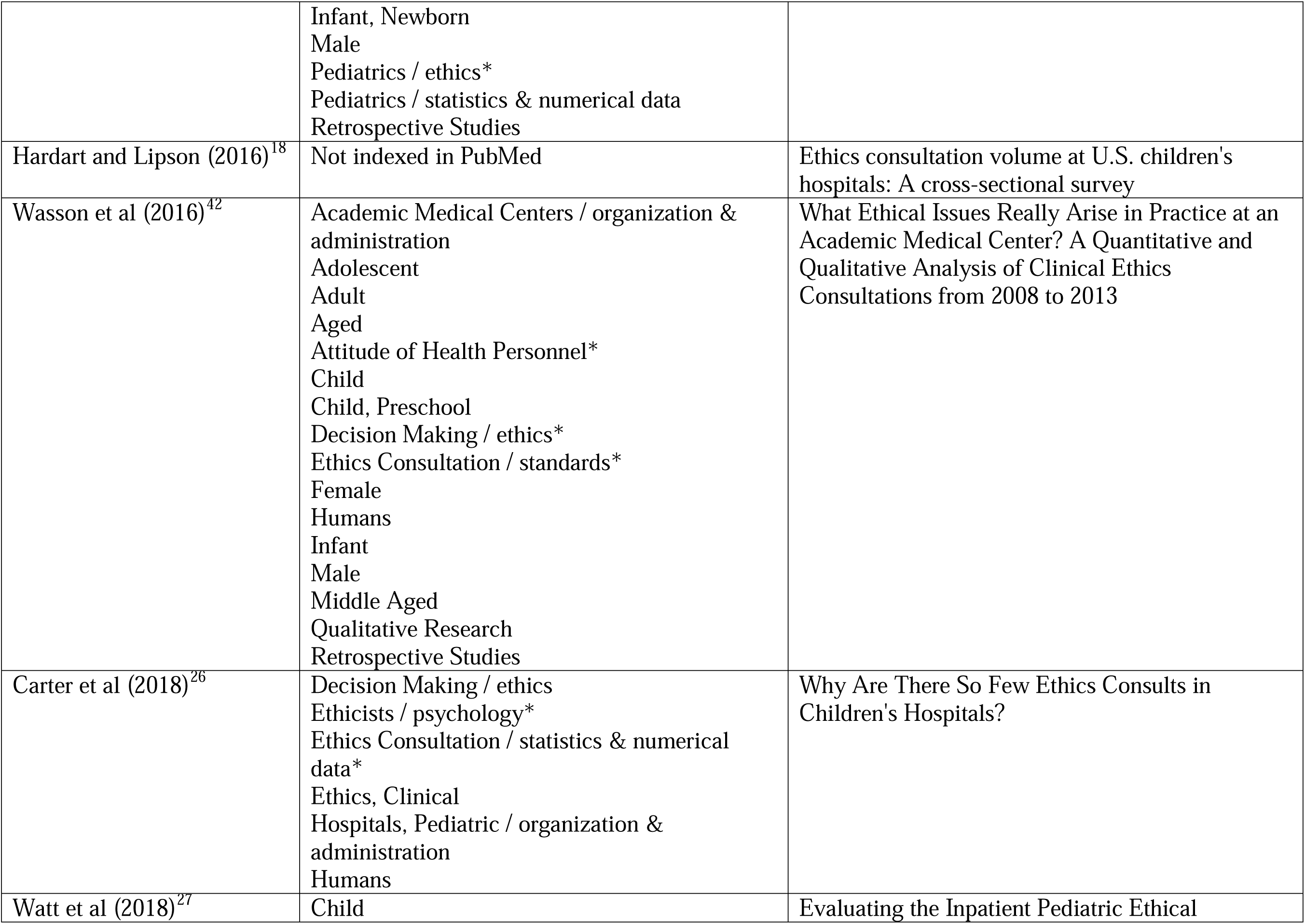
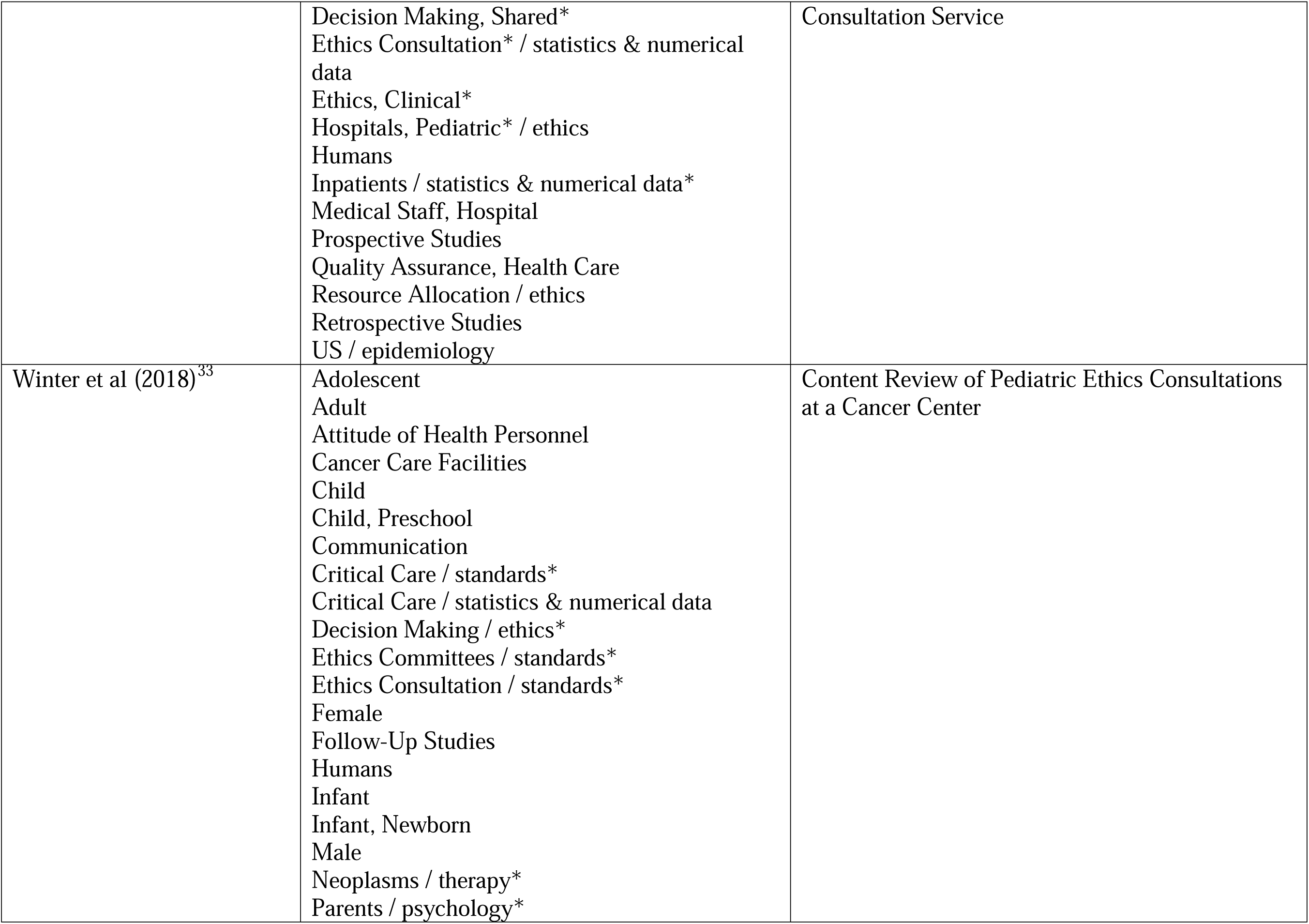
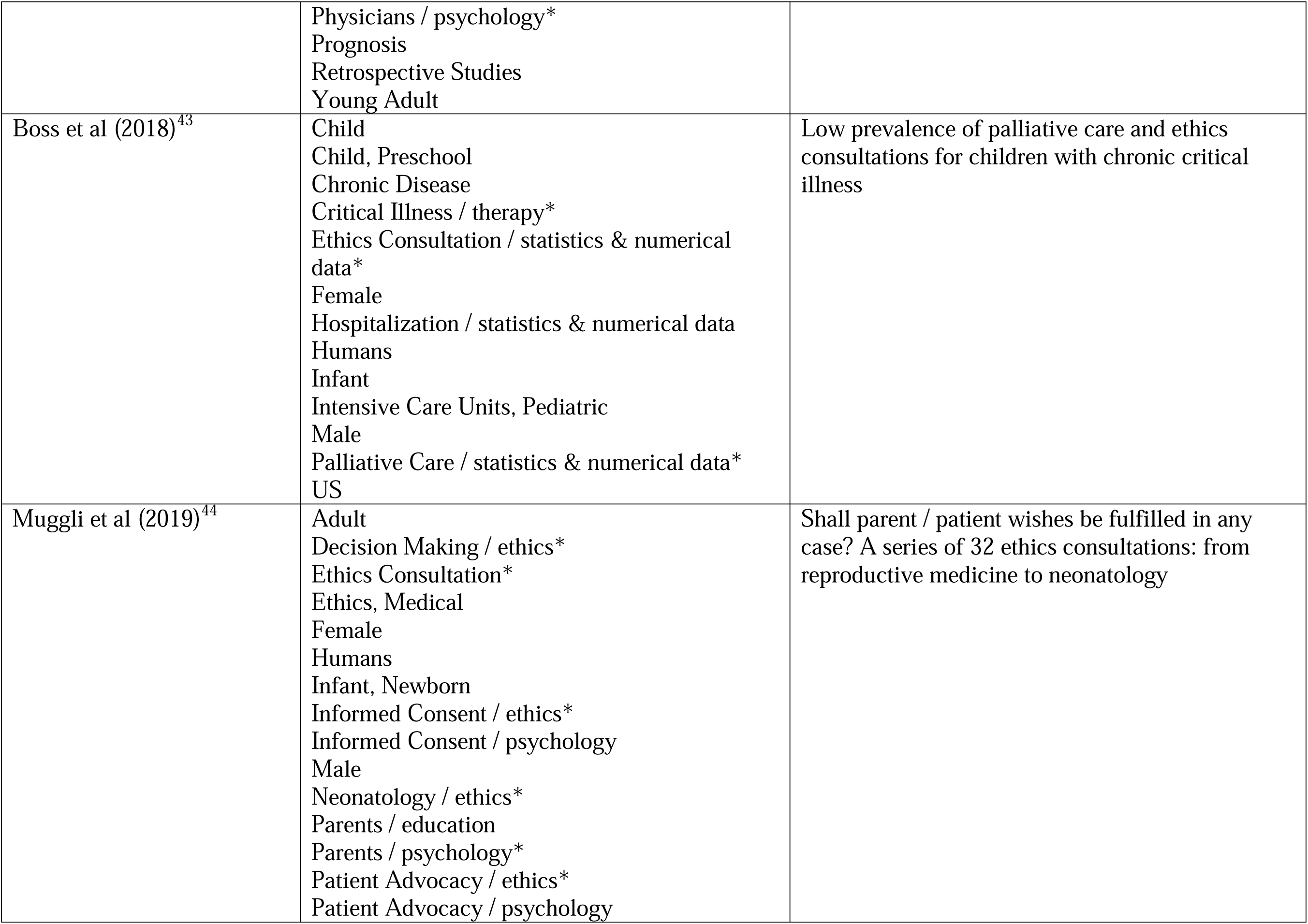
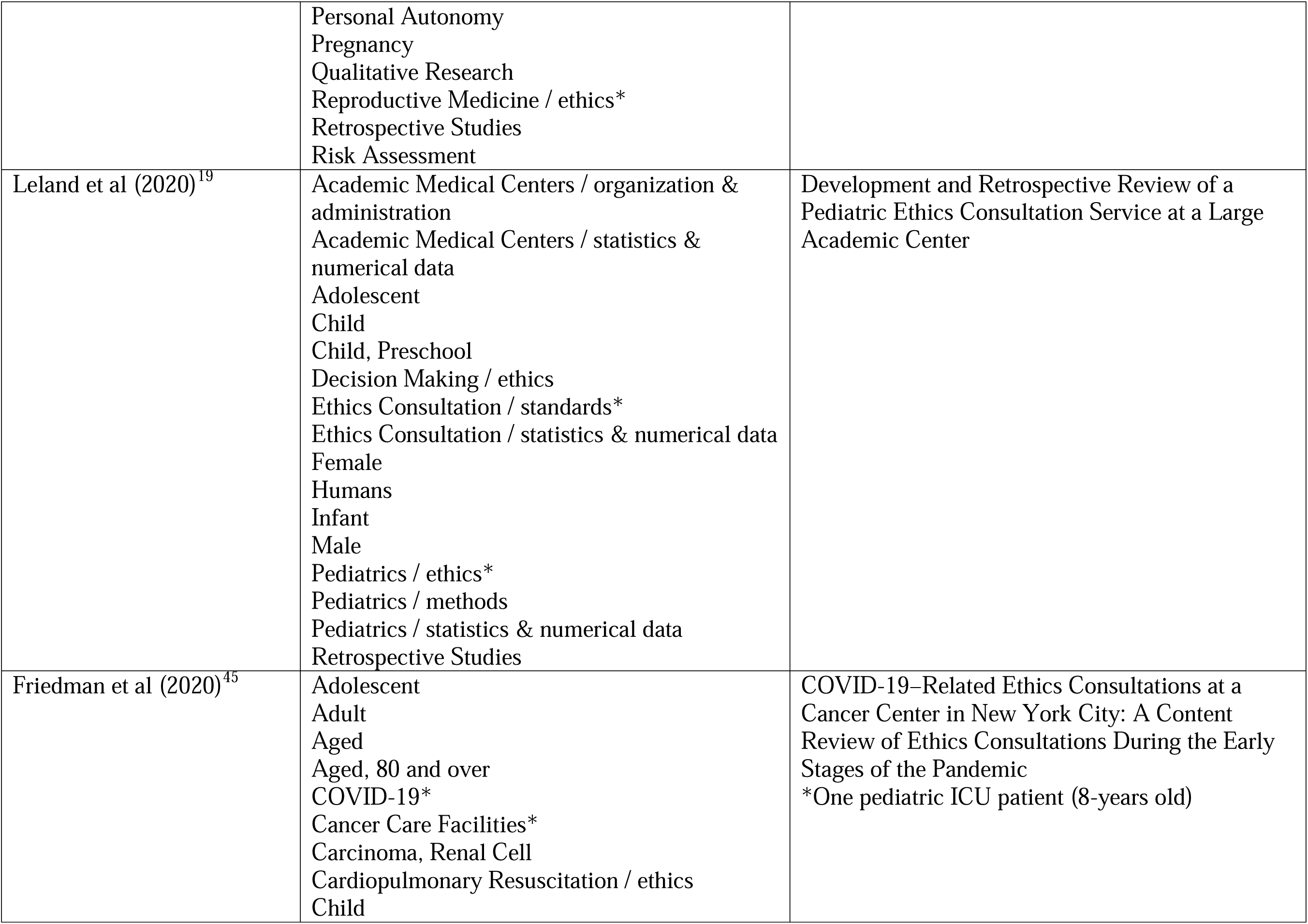
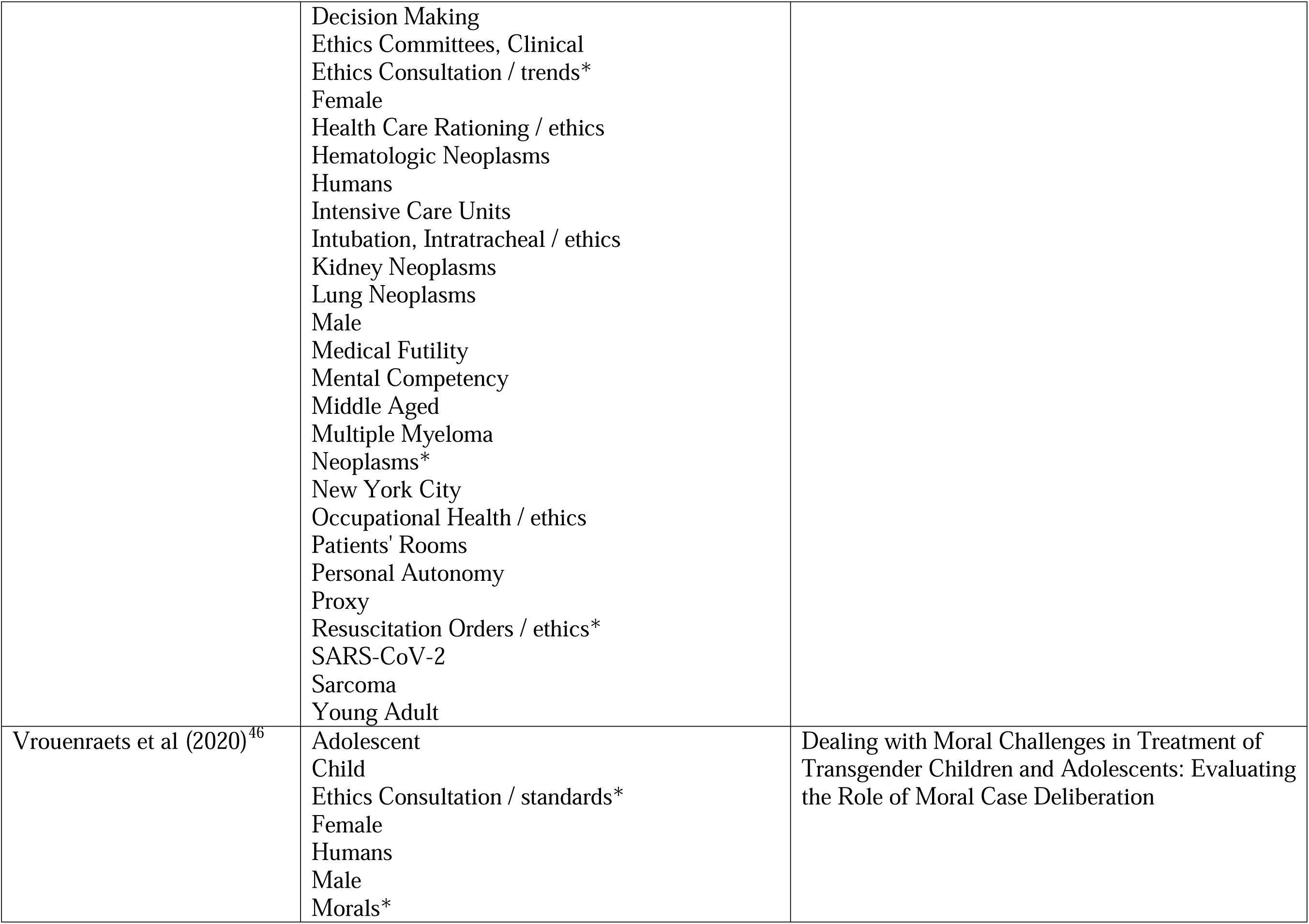
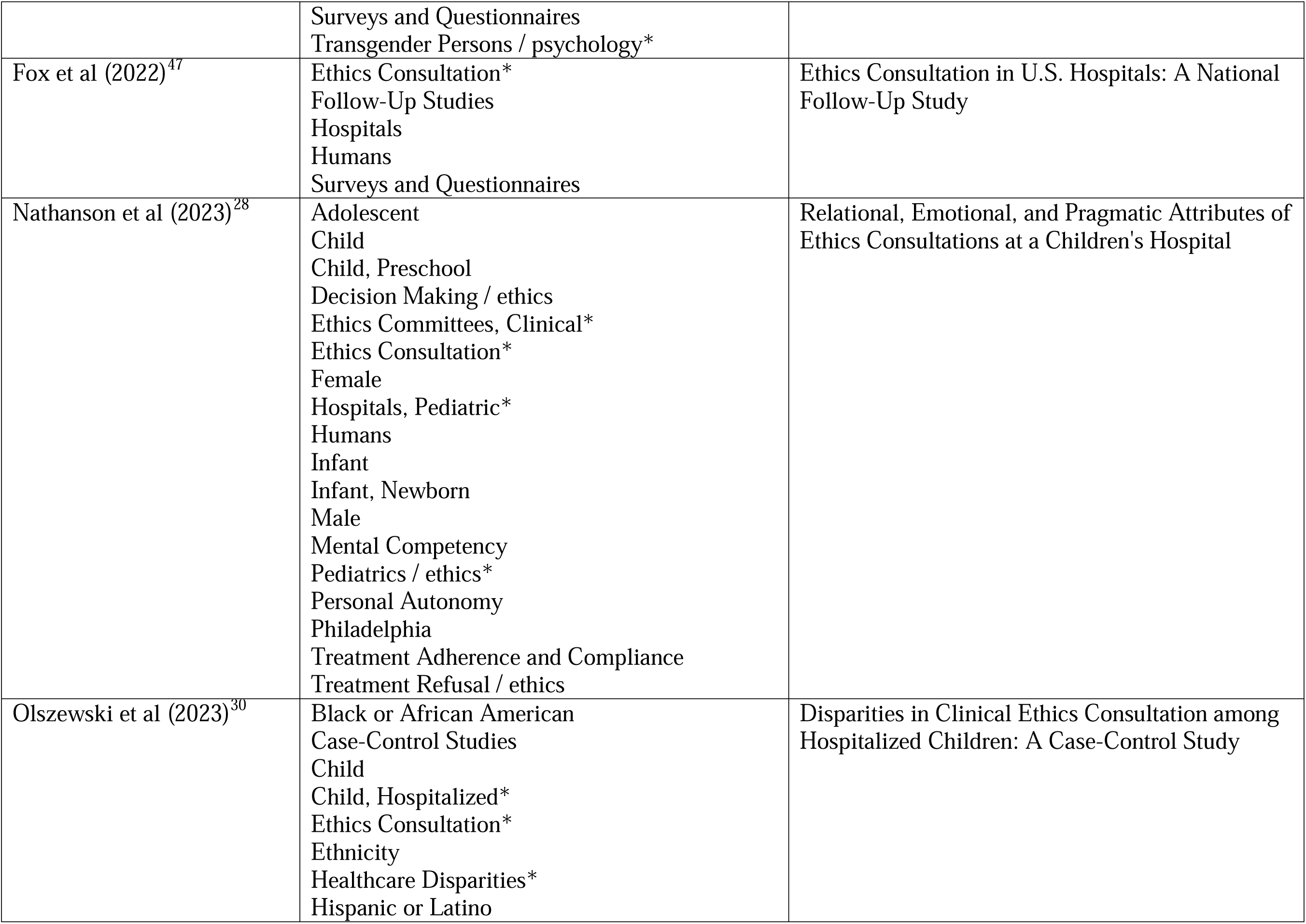
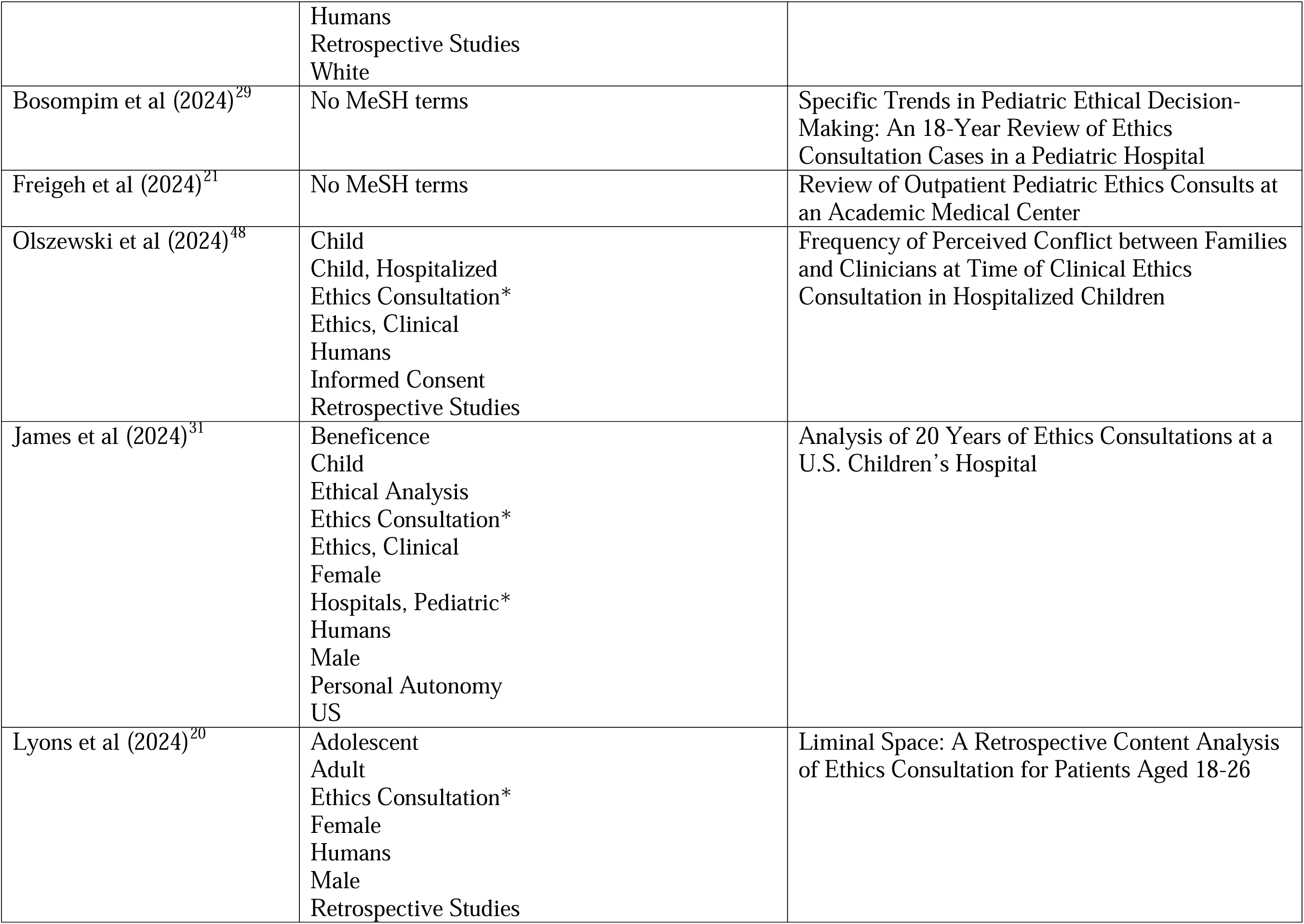
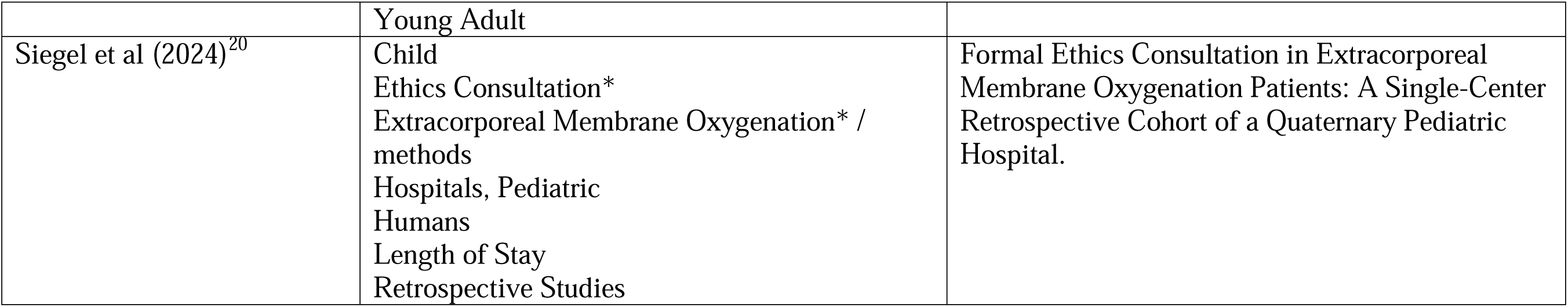
MeSH terms and title/text words from relevant studies (n=32) used to generate search strategy.

| Author (publication date) | MeSH terms | Title / Words |
| --- | --- | --- |
| La Puma (1987)<sup>38</sup> | Adolescent<br>Adult<br>Aged<br>Aged, 80 and over<br>Bioethical Issues<br>Bioethics*<br>Child<br>Ethicists*<br>Ethics Committees, Clinical*<br>Ethics Consultation*<br>Ethics, Medical*<br>Female<br>Humans<br>Life Support Care<br>Male<br>Middle Aged<br>Referral and Consultation*<br>US<br>Withholding Treatment | Consultations in clinical ethics--issues and questions in 27 cases. |
| La Puma (1992)<sup>39</sup> | Adolescent<br>Adult<br>Advance Directives<br>Aged<br>Aged, 80 and over<br>Attitude of Health Personnel<br>Bioethical Issues<br>Child<br>Child, Preschool<br>Ethicists*<br>Ethics Consultation*<br>Ethics, Clinical* | Community hospital ethics consultation: evaluation and comparison with a university hospital service |
|  | Ethics, Medical*<br>Evaluation Studies as Topic<br>Female<br>Hospitals, Community / standards*<br>Hospitals, University / standards*<br>Humans<br>Internship and Residency<br>Interprofessional Relations<br>Interviews as Topic<br>Male<br>Medical Staff, Hospital<br>Mental Competency<br>Middle Aged<br>Physician-Patient Relations<br>Prospective Studies<br>Referral and Consultation* |  |
| Orr and Perkin (1994)<sup>22</sup> | Adolescent<br>California<br>Child<br>Child Custody / legislation & jurisprudence*<br>Child, Preschool<br>Dissent and Disputes*<br>Ethicists*<br>Ethics Committees<br>Ethics Consultation*<br>Ethics, Clinical<br>Ethics, Medical*<br>Female<br>Group Processes*<br>Hospital Bed Capacity, 500 and over<br>Humans<br>Infant<br>Infant, Newborn | Clinical Ethics Consultations with Children |
|  | Interprofessional Relations<br>Legal Guardians*<br>Male<br>Patient Advocacy / legislation & jurisprudence<br>Patient Care Team / legislation & jurisprudence<br>Physician-Patient Relations*<br>Professional Role<br>Professional-Family Relations<br>Referral and Consultation / legislation & jurisprudence<br>Referral and Consultation / statistics & numerical data<br>Withholding Treatment |  |
| Yen Schneiderman (1999)<sup>15</sup> | Adolescent<br>Child<br>Child, Preschool<br>Communication<br>Ethics, Medical*<br>Evaluation Studies as Topic<br>Family*<br>Humans<br>Infant<br>Infant, Newborn<br>Male<br>Patients<br>Pediatrics / methods*<br>Personal Satisfaction<br>Physicians*<br>Referral and Consultation* / standards<br>Retrospective Studies<br>Social Work* | Impact of pediatric ethics consultations on patients, families, social workers, and physicians |
| Mitchell and Truog (2000)<sup>50</sup> | Adolescent<br>Boston | From the Files of a Pediatric Ethics Committee |
|  | Child<br>Child, Preschool<br>Decision Making<br>Ethics Committees, Clinical*<br>Ethics Consultation*<br>Ethics, Clinical*<br>Ethics, Institutional<br>Ethics, Medical*<br>Group Processes<br>Humans<br>Infant<br>Infant, Newborn<br>Parental Consent<br>Pediatrics / standards*<br>Professional-Family Relations |  |
| McGee et al (2001)<sup>16</sup> | Academic Medical Centers / standards*<br>Ethicists*<br>Ethics Consultation*<br>Ethics, Clinical*<br>Ethics, Medical*<br>Hospital Units<br>Humans<br>North Carolina<br>Patient Care Team<br>Philadelphia<br>Professional Practice*<br>Professional Role*<br>Referral and Consultation*<br>Role<br>Social Responsibility | Paradigms for clinical ethics consultation practice |
| Fox et al (2000)<sup>41</sup> | Ethics Committees, Clinical / statistics & numerical data*<br>Ethics Consultation / statistics & numerical | Ethics consultation in US hospitals: a national survey |
|  | data*<br>Hospital Bed Capacity<br>Humans<br>Patient Rights*<br>Surveys and Questionnaires<br>US |  |
| Opel et al (2009)<sup>23</sup> | Decision Making / ethics*<br>Ethics Consultation*<br>Female<br>Humans<br>Male<br>Pediatrics / ethics* | Pediatric clinical ethics consultations at an academic medical center: does one size fit all? |
| Streuli et al (2014)<sup>24</sup> | Adolescent<br>Child<br>Child, Preschool<br>Decision Making<br>Ethics Committees<br>Ethics Consultation*<br>Ethics, Clinical*<br>Female<br>Hospitals, Pediatric*<br>Hospitals, Teaching*<br>Humans<br>Infant<br>Infant, Newborn<br>Male<br>Pediatrics / ethics*<br>Quality-Adjusted Life Years<br>Retrospective Studies<br>Switzerland | Five-year experience of clinical ethics consultations in a pediatric teaching hospital |
| Johnson et al (2015)<sup>32</sup> | Academic Medical Centers<br>Adolescent<br>Adult | Ethics consultation in pediatrics: long-term experience from a pediatric oncology center |
|  | Child<br>Child, Preschool<br>Decision Making / ethics*<br>Ethics Committees, Research<br>Ethics Consultation*<br>Female<br>Hospitals, Pediatric<br>Humans<br>Infant<br>Male<br>Moral Obligations<br>Negotiating<br>Palliative Care / ethics<br>Parents<br>Pediatrics / ethics*<br>Personal Autonomy<br>Retrospective Studies<br>Risk Assessment<br>Tennessee |  |
| Henricksen et al (2015)<sup>17</sup> | Decision Making / ethics*<br>Ethics Consultation*<br>Female<br>Humans<br>Male<br>Pediatrics / ethics* | Pediatric clinical ethics consultations at an academic medical center: does one size fit all? |
| Thomas et al (2015)<sup>25</sup> | Adolescent<br>Child<br>Child, Preschool<br>Ethics Consultation / statistics & numerical data*<br>Female<br>Humans<br>Infant | Not just little adults: a review of 102 paediatric ethics consultations |
|  | Infant, Newborn<br>Male<br>Pediatrics / ethics*<br>Pediatrics / statistics & numerical data<br>Retrospective Studies |  |
| Hardart and Lipson (2016)<sup>18</sup> | Not indexed in PubMed | Ethics consultation volume at U.S. children's hospitals: A cross-sectional survey |
| Wasson et al (2016)<sup>42</sup> | Academic Medical Centers / organization & administration<br>Adolescent<br>Adult<br>Aged<br>Attitude of Health Personnel*<br>Child<br>Child, Preschool<br>Decision Making / ethics*<br>Ethics Consultation / standards*<br>Female<br>Humans<br>Infant<br>Male<br>Middle Aged<br>Qualitative Research<br>Retrospective Studies | What Ethical Issues Really Arise in Practice at an Academic Medical Center? A Quantitative and Qualitative Analysis of Clinical Ethics Consultations from 2008 to 2013 |
| Carter et al (2018)<sup>26</sup> | Decision Making / ethics<br>Ethicists / psychology*<br>Ethics Consultation / statistics & numerical data*<br>Ethics, Clinical<br>Hospitals, Pediatric / organization & administration<br>Humans | Why Are There So Few Ethics Consults in Children's Hospitals? |
| Watt et al (2018)<sup>27</sup> | Child | Evaluating the Inpatient Pediatric Ethical |
|  | Decision Making, Shared*<br>Ethics Consultation* / statistics & numerical data<br>Ethics, Clinical*<br>Hospitals, Pediatric* / ethics<br>Humans<br>Inpatients / statistics & numerical data*<br>Medical Staff, Hospital<br>Prospective Studies<br>Quality Assurance, Health Care<br>Resource Allocation / ethics<br>Retrospective Studies<br>US / epidemiology | Consultation Service |
| Winter et al (2018)<sup>33</sup> | Adolescent<br>Adult<br>Attitude of Health Personnel<br>Cancer Care Facilities<br>Child<br>Child, Preschool<br>Communication<br>Critical Care / standards*<br>Critical Care / statistics & numerical data<br>Decision Making / ethics*<br>Ethics Committees / standards*<br>Ethics Consultation / standards*<br>Female<br>Follow-Up Studies<br>Humans<br>Infant<br>Infant, Newborn<br>Male<br>Neoplasms / therapy*<br>Parents / psychology* | Content Review of Pediatric Ethics Consultations at a Cancer Center |
|  | Physicians / psychology*<br>Prognosis<br>Retrospective Studies<br>Young Adult |  |
| Boss et al (2018)<sup>43</sup> | Child<br>Child, Preschool<br>Chronic Disease<br>Critical Illness / therapy*<br>Ethics Consultation / statistics & numerical data*<br>Female<br>Hospitalization / statistics & numerical data<br>Humans<br>Infant<br>Intensive Care Units, Pediatric<br>Male<br>Palliative Care / statistics & numerical data*<br>US | Low prevalence of palliative care and ethics consultations for children with chronic critical illness |
| Muggli et al (2019)<sup>44</sup> | Adult<br>Decision Making / ethics*<br>Ethics Consultation*<br>Ethics, Medical<br>Female<br>Humans<br>Infant, Newborn<br>Informed Consent / ethics*<br>Informed Consent / psychology<br>Male<br>Neonatology / ethics*<br>Parents / education<br>Parents / psychology*<br>Patient Advocacy / ethics*<br>Patient Advocacy / psychology | Shall parent / patient wishes be fulfilled in any case? A series of 32 ethics consultations: from reproductive medicine to neonatology |
|  | Personal Autonomy<br>Pregnancy<br>Qualitative Research<br>Reproductive Medicine / ethics*<br>Retrospective Studies<br>Risk Assessment |  |
| Leland et al (2020)<sup>19</sup> | Academic Medical Centers / organization & administration<br>Academic Medical Centers / statistics & numerical data<br>Adolescent<br>Child<br>Child, Preschool<br>Decision Making / ethics<br>Ethics Consultation / standards*<br>Ethics Consultation / statistics & numerical data<br>Female<br>Humans<br>Infant<br>Male<br>Pediatrics / ethics*<br>Pediatrics / methods<br>Pediatrics / statistics & numerical data<br>Retrospective Studies | Development and Retrospective Review of a Pediatric Ethics Consultation Service at a Large Academic Center |
| Friedman et al (2020)<sup>45</sup> | Adolescent<br>Adult<br>Aged<br>Aged, 80 and over<br>COVID-19*<br>Cancer Care Facilities*<br>Carcinoma, Renal Cell<br>Cardiopulmonary Resuscitation / ethics<br>Child | COVID-19–Related Ethics Consultations at a Cancer Center in New York City: A Content Review of Ethics Consultations During the Early Stages of the Pandemic<br>*One pediatric ICU patient (8-years old) |
|  | Decision Making<br>Ethics Committees, Clinical<br>Ethics Consultation / trends*<br>Female<br>Health Care Rationing / ethics<br>Hematologic Neoplasms<br>Humans<br>Intensive Care Units<br>Intubation, Intratracheal / ethics<br>Kidney Neoplasms<br>Lung Neoplasms<br>Male<br>Medical Futility<br>Mental Competency<br>Middle Aged<br>Multiple Myeloma<br>Neoplasms*<br>New York City<br>Occupational Health / ethics<br>Patients' Rooms<br>Personal Autonomy<br>Proxy<br>Resuscitation Orders / ethics*<br>SARS-CoV-2<br>Sarcoma<br>Young Adult |  |
| Vrouenraets et al (2020)<sup>46</sup> | Adolescent<br>Child<br>Ethics Consultation / standards*<br>Female<br>Humans<br>Male<br>Morals* | Dealing with Moral Challenges in Treatment of Transgender Children and Adolescents: Evaluating the Role of Moral Case Deliberation |
|  | Surveys and Questionnaires<br>Transgender Persons / psychology* |  |
| Fox et al (2022)<sup>47</sup> | Ethics Consultation*<br>Follow-Up Studies<br>Hospitals<br>Humans<br>Surveys and Questionnaires | Ethics Consultation in U.S. Hospitals: A National Follow-Up Study |
| Nathanson et al (2023)<sup>28</sup> | Adolescent<br>Child<br>Child, Preschool<br>Decision Making / ethics<br>Ethics Committees, Clinical*<br>Ethics Consultation*<br>Female<br>Hospitals, Pediatric*<br>Humans<br>Infant<br>Infant, Newborn<br>Male<br>Mental Competency<br>Pediatrics / ethics*<br>Personal Autonomy<br>Philadelphia<br>Treatment Adherence and Compliance<br>Treatment Refusal / ethics | Relational, Emotional, and Pragmatic Attributes of Ethics Consultations at a Children's Hospital |
| Olszewski et al (2023)<sup>30</sup> | Black or African American<br>Case-Control Studies<br>Child<br>Child, Hospitalized*<br>Ethics Consultation*<br>Ethnicity<br>Healthcare Disparities*<br>Hispanic or Latino | Disparities in Clinical Ethics Consultation among Hospitalized Children: A Case-Control Study |
|  | Humans<br>Retrospective Studies<br>White |  |
| Bosompim et al (2024)<sup>29</sup> | No MeSH terms | Specific Trends in Pediatric Ethical Decision-Making: An 18-Year Review of Ethics Consultation Cases in a Pediatric Hospital |
| Freigeh et al (2024)<sup>21</sup> | No MeSH terms | Review of Outpatient Pediatric Ethics Consults at an Academic Medical Center |
| Olszewski et al (2024)<sup>48</sup> | Child<br>Child, Hospitalized<br>Ethics Consultation*<br>Ethics, Clinical<br>Humans<br>Informed Consent<br>Retrospective Studies | Frequency of Perceived Conflict between Families and Clinicians at Time of Clinical Ethics Consultation in Hospitalized Children |
| James et al (2024)<sup>31</sup> | Beneficence<br>Child<br>Ethical Analysis<br>Ethics Consultation*<br>Ethics, Clinical<br>Female<br>Hospitals, Pediatric*<br>Humans<br>Male<br>Personal Autonomy<br>US | Analysis of 20 Years of Ethics Consultations at a U.S. Children's Hospital |
| Lyons et al (2024)<sup>20</sup> | Adolescent<br>Adult<br>Ethics Consultation*<br>Female<br>Humans<br>Male<br>Retrospective Studies | Liminal Space: A Retrospective Content Analysis of Ethics Consultation for Patients Aged 18-26 |
|  | Young Adult |  |
| Siegel et al (2024)<sup>20</sup> | Child<br>Ethics Consultation*<br>Extracorporeal Membrane Oxygenation* /<br>methods<br>Hospitals, Pediatric<br>Humans<br>Length of Stay<br>Retrospective Studies | Formal Ethics Consultation in Extracorporeal<br>Membrane Oxygenation Patients: A Single-Center<br>Retrospective Cohort of a Quaternary Pediatric<br>Hospital. |

